# Non-invasive vagus nerve stimulation conditions increased invigoration and wanting in depression

**DOI:** 10.1101/2023.09.28.23296284

**Authors:** Magdalena Ferstl, Anne Kühnel, Johannes Klaus, Wy Ming Lin, Nils B. Kroemer

## Abstract

**Background:** Major depressive disorder (MDD) is often marked by impaired motivation and reward processing, known as anhedonia. Many patients do not respond to first-line treatments, and improvements in motivation can be slow, creating an urgent need for rapid interventions. Recently, we demonstrated that transcutaneous auricular vagus nerve stimulation (taVNS) acutely boosts effort invigoration in healthy participants, but its effects on depression remain unclear.

**Objective:** To assess the impact of taVNS on effort invigoration and maintenance in a sample that includes patients with MDD, evaluating the generalizability of our findings.

**Methods:** We used a single-blind, randomized crossover design in 30 patients with MDD and 29 matched (age, sex, and BMI) healthy control participants (HCP).

**Results:** Consistent with prior findings, taVNS increased effort invigoration for rewards in both groups during Session 1 (*p*=.040), particularly for less wanted rewards in HCP (*p*_boot_<.001). However, invigoration remained elevated in all participants, and no acute changes were observed in Session 2 (Δinvigoration=3.52, *p*=.093). Crucially, throughout Session 1, we found taVNS-induced increases in effort invigoration (*p*_boot_ =.008) and wanting (*p*_boot_=.010) in patients with MDD, with gains in wanting maintained across sessions (Δwanting=0.06, *p*=.97).

**Conclusions:** Our study replicates the invigorating effects of taVNS in Session 1 and reveals its generalizability to depression. Furthermore, we expand upon previous research by showing taVNS-induced conditioning effects on invigoration and wanting within Session 1 in patients that were largely sustained. While enduring motivational improvements present challenges for crossover designs, they are highly desirable in interventions and warrant further follow-up research.

**Graphical abstract:** 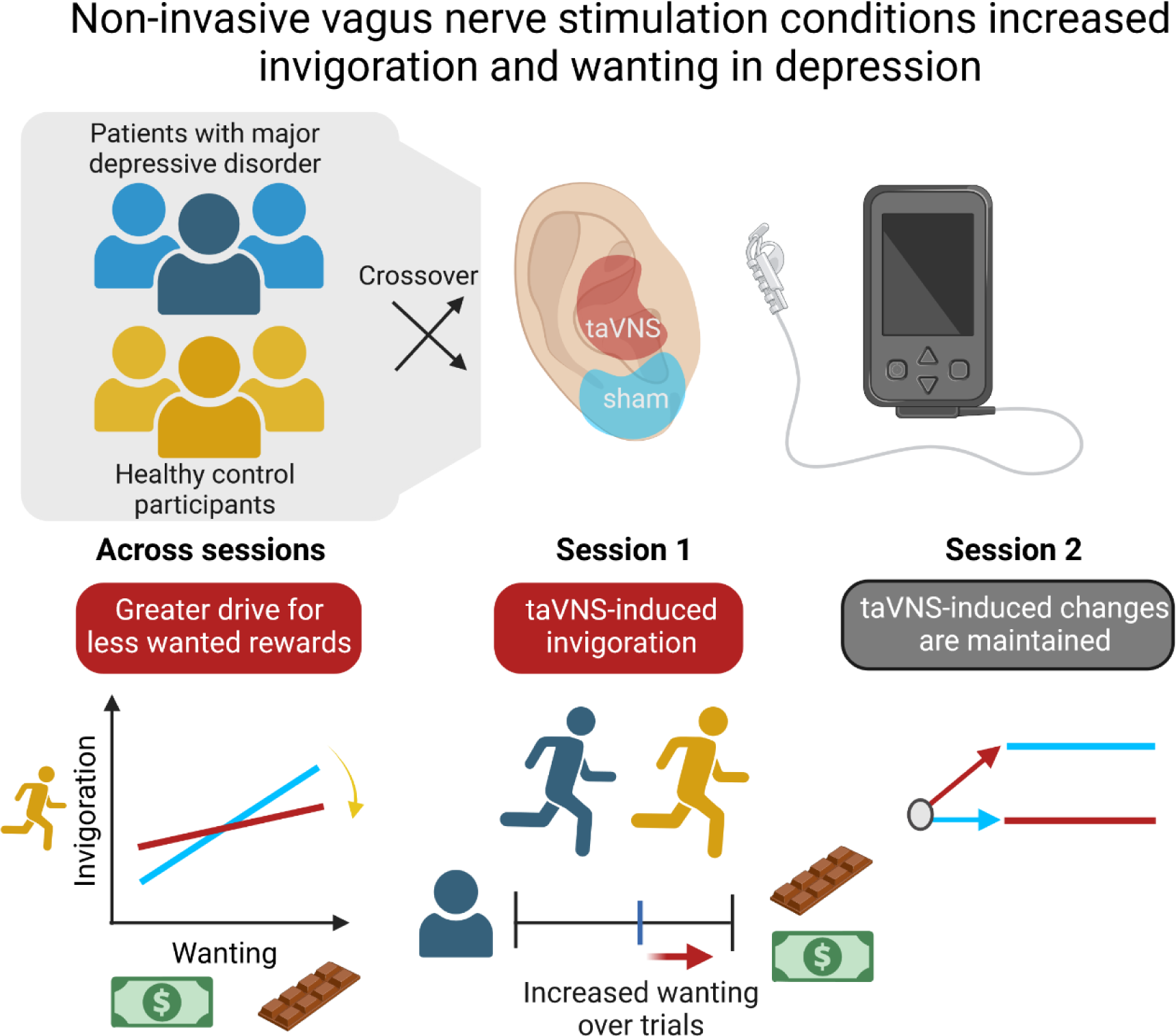

## Introduction

While we often experience temporary low moods and motivation, persistent negative feelings may signify depressive episodes instead. With over 280 million people affected globally, major depressive disorder (MDD) is a leading cause of disability ^1, 2^. The prevalence of MDD highlights the need for improved treatment options. Particularly, somatic symptoms ^3^ and anhedonia are associated with greater severity and persistence of MDD ^4, 5^ and they respond worse to conventional treatments ^4, 6^. Recent hypotheses implicate aberrant interoceptive signaling as a potential cause in the etiology of somatic and motivational symptoms ^7–9^. The required communication between peripheral organs and the brain is channeled through the vagus nerve and converges on the nucleus of the solitary tract in the brainstem to modulate motivational circuits ^10^. Consequently, vagal afferents tune motivated behavior ^11, 12^ such as food seeking ^13, 14^ and response vigor ^15^. While preliminary evidence has shown the potential of transcutaneous vagus nerve stimulation (taVNS) to affect motivated behavior and brain–body signaling in healthy participants ^16, 17^, its potential for improving motivational deficits in MDD has not yet been established.

Anhedonia is a cardinal symptom of depression and involves diminished reward anticipation and effort exertion ^18–20^. Although rewards typically boost motivation ^21–23^, this function appears impaired in patients with MDD ^20, 24^. In addition to struggling to adapt their behavior to reward magnitude, these patients often experience increased exhaustion after effort tasks ^21, 24, 25^. Since this motivation deficit worsens with symptom severity and correlates with a poorer prognosis ^20, 26^, there is an unmet need for rapid, targeted treatments addressing motivational alterations in MDD.

To treat symptoms of anhedonia and boost mood in people with depression, pharmacotherapy with selective serotonin reuptake inhibitors (SSRIs) is the first line of treatment ^27, 28^. On the behavioral level, SSRIs have been shown to increase effort expenditure for rewards ^29^, a behavioral proxy of motivational facets of anhedonia ^30^. Still, antidepressant medication only improves reward-related symptoms in a subset of patients ^31^ and anhedonia is less responsive to SSRI treatments compared to mood-related symptoms ^32^. Moreover, the antidepressant effects of SSRIs only materialize after several weeks of use ^33–36^. Hence, adjunct treatments, such as vagus nerve stimulation (VNS), have been proposed to improve symptoms more rapidly and increase overall response rates ^37, 38^. Indeed, studies have shown antidepressant effects of invasive VNS ^39–41^ and non-invasive VNS ^42–45^. In addition to motivational effects, non-invasive VNS has been shown to acutely counteract anxiety-related mechanisms and symptoms ^46, 47^. Since non-invasive VNS is well-tolerated ^48–50^, it could be used as an adjunct treatment for motivational or somatic symptoms.

Despite the promising long-term effectiveness of VNS ^51–53^, the acute effects on motivational symptoms remain largely elusive ^50^. In healthy participants, we have recently shown that taVNS boosts the drive to work for rewards ^16^ and ameliorates the dampened mood after effortful tasks ^54^. Since taVNS-induced improvements in motivation were stronger for participants with low baseline mood, it might be a promising technique to rapidly increase motivational drive in patients with MDD as well. To close this gap, we investigated taVNS-induced changes in the motivation to work for rewards^16^ using a single-blind crossover design (taVNS vs. sham) in participants with MDD as well as matched healthy control participants (HCP). Based on preclinical findings and our previous results, we expected taVNS to acutely boost motivation, generalizing to patients with MDD. In accordance with this hypothesis, we observed taVNS-induced increases in invigoration during the first session, whereas there was no acute effect of taVNS in the second session. Intriguingly, we observed conditioning effects in patients with MDD during the first session that were largely maintained, pointing to a potentially longer-lasting improvement elicited by taVNS.

## Methods

### Participants

We included 31 participants with MDD as well as 34 HCP in the study who completed two sessions: one with right-sided taVNS and the other with sham stimulation (order randomized). However, data had to be excluded from the analysis due to dropout (n=2), technical issues (n=3), or medication (1 HCP), leading to a final sample size of N=59 (i.e., n=29 HCP and n=30 with MDD, Table 1). To be included, participants went through a screening protocol to ensure they were physically healthy, 18 to 65 years old, and within a normal or overweight range of their body mass index (BMI, 18.5-30 kg/m^2^). Participants with MDD had to fulfill the criteria for MDD ^1^ within the last 12 months and had to have a current BDI-II score ≥14 ^55^. In contrast, HCPs had no history of depression. We only included participants without other mental comorbidities apart from anxiety disorders (in HCP only specific phobias) and tobacco use disorder ^56^. Participants provided written informed consent at the beginning of Session 1 and received either monetary compensation (32€ fixed amount) or course credit for their participation after completing the second session. Moreover, they received money and snacks depending on their performance during the tasks. The study protocol was registered at clinicaltrials.gov (NCT05120336). The study was approved by the ethics committee of the medical faculty of the university of Tübingen and conducted in accordance with the ethical code of the World Medical Association (Declaration of Helsinki).

**Table 1:** Characteristics of the sample. Values reported are mean (SD) for continuous variables and n (%) for categorical variables. ^1^Pearson’s Chi-squared test; ^2^Wilcoxon rank sum test; ^3^Fisher’s exact test. taVNS = transcutaneous vagus nerve stimulation, SSRI = selective serotonin reuptake inhibitor, SNRI = selective noradrenaline reuptake inhibitor, BMI = Body mass index, BDI = Beck Depression inventory, SHAPS = Snaith-Hamilton Pleasure Scale.

| Variable | N | Overall, N=59 | HCP, N=29 | MDD, N=30 | p-value<sup>2</sup> |
| --- | --- | --- | --- | --- | --- |
| **Sex** | 59 |  |  |  | 0.71<sup>1</sup> |
| male |  | 19 (32%) | 10 (34%) | 9 (30%) |  |
| female |  | 40 (68%) | 19 (66%) | 21 (70%) |  |
| **Age** | 59 | 34.2 (13.5) | 37.1 (14.5) | 31.4 (12.1) | 0.12<sup>2</sup> |
| **Stimulation Session 1** | 59 |  |  |  | 0.70<sup>1</sup> |
| Sham |  | 30 (51%) | 14 (48%) | 16 (53%) |  |
| taVNS |  | 29 (49%) | 15 (52%) | 14 (47%) |  |
| **Comorbidities** |  |  |  |  |  |
| Anxiety | 59 | 13 (22%) | 5 (17%) | 8 (27%) | 0.38<sup>1</sup> |
| Trauma-related | 59 | 2 (3.4%) | 0 (0%) | 2 (6.7%) | 0.49<sup>3</sup> |
| Substance dependence (Tobacco) | 59 | 5 (8.5%) | 3 (10%) | 2 (6.7%) | 0.67<sup>3</sup> |
| Personality disorders | 59 | 1 (1.7%) | 0 (0%) | 1 (3.3%) | >0.99<sup>3</sup> |
| **Medication** |  |  |  |  |  |
| SSRI | 59 | 8 (14%) | 0 (0%) | 8 (27%) | 0.005<sup>3</sup> |
| SNRI | 59 | 4 (6.8%) | 0 (0%) | 4 (13%) | 0.11<sup>3</sup> |
| Other | 59 | 3 (5.1%) | 0 (0%) | 3 (10%) | 0.24<sup>3</sup> |
| Tetracyclic | 59 | 3 (5.1%) | 0 (0%) | 3 (10%) | 0.24<sup>3</sup> |
| Atypical antipsychotics | 59 | 2 (3.4%) | 0 (0%) | 2 (6.7%) | 0.49<sup>3</sup> |
| Tricyclic | 59 | 1 (1.7%) | 0 (0%) | 1 (3.3%) | >0.99<sup>3</sup> |
| **BMI [kg/m<sup>2</sup>]** | 59 | 24.3 (3.4) | 24.0 (3.4) | 24.6 (3.5) | 0.42<sup>2</sup> |
| **BDI** | 59 | 13.1 (11.3) | 4.1 (3.7) | 21.7 (9.2) | <0.001<sup>2</sup> |
| Minimal to mild |  |  |  | 14 (48%) |  |
| Moderate |  |  |  | 7 (24%) |  |
| Severe |  |  |  | 8 (29%) |  |
| **SHAPS** | 59 | 2.5 (2.9) | 0.9 (2.5) | 4.1 (2.4) | <0.001<sup>2</sup> |

### Experimental procedure

Experimental sessions were conducted in a randomized, single-blind crossover design that was adapted with minor modifications from our previous studies (see ^16, 54^, Figure 1). Before the first session, participants were asked to answer several questionnaires at home, including the Beck Depression Inventory-II (BDI-II; ^55^), Snaith Hamilton Pleasure Scale (SHAPS; ^57^) and behavioral inhibition/activation scales (BIS/BAS; ^58^) to assess reward processing and quantify depressive symptoms.

**Figure 1.**
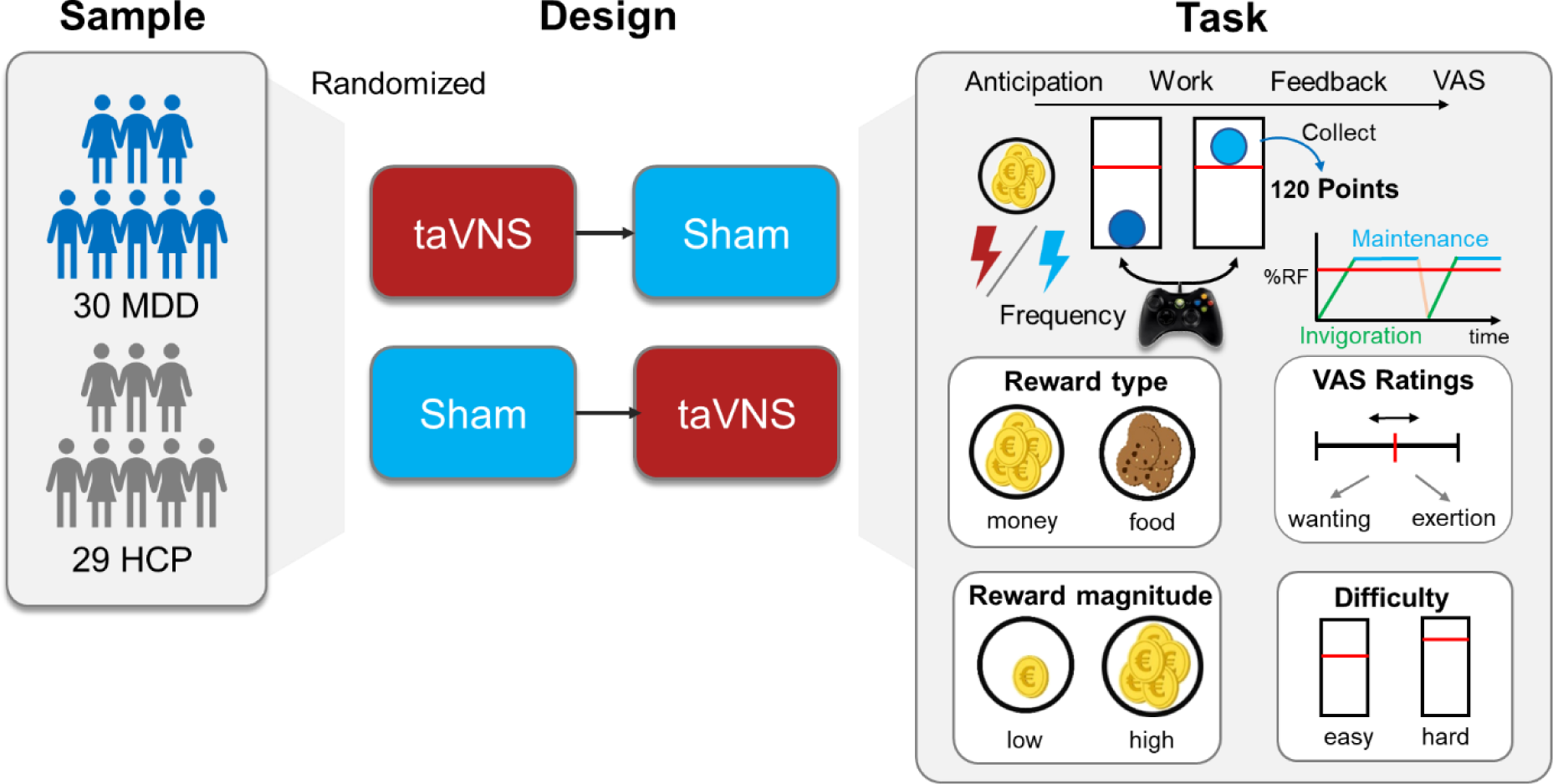
Experimental procedure and overview of the effort allocation task (EAT). Participants (29 healthy control participants, HCP, and 30 with major depressive disorder, MDD) completed two experimental sessions, one with active transcutaneous auricular vagus nerve stimulation (taVNS) and one with sham stimulation (randomized crossover). Within each session, participants complete tasks during stimulation. In the EAT, participants have to repeatedly press a button on an Xbox controller to elevate a ball above a red difficulty line to collect food and monetary reward points. Trials differ in their reward magnitude, reward type, and difficulty. After each trial, participants rated how much they wanted to receive the reward and how much they exerted themselves using visual analog scales (VAS). RF = relative frequency (to the maximal frequency of the participant during the training)

Participants were asked to fast 3-5h before each session (M_fast_ = 4.05h ± 2.12h) so that they felt neither full nor hungry. Sessions started at approximately the same time of the day (±1h) and lasted about 2.5h. In Session 1, participants provided written informed consent. After measuring physiological and anthropometric parameters (e.g., pulse and weight), we recorded their preceding food and drink intake. Participants were allowed to drink water during the whole session. Next, participants reported their current mood state based on the Positive And Negative Affect Scale (PANAS; Watson ^59^) using visual analog scales (VAS). To practice the effort task and calibrate the individual maximum frequency of button presses, we included a training run of the effort allocation task (EAT). Then, the stimulation electrode of the NEMOS® tVNS device (cerbomed GmbH, Erlangen, Germany) was attached and secured with surgical tape on the right ear, either at the cymba conchae for taVNS or at the ear lobe for sham. For each session, we individually adjusted the stimulation amplitude by slowly increasing it in 0.1-0.2 mA steps until the participants’ sensation (assessed with VAS) was rated as a “mild pricking” and below the pain threshold ^16, 60, 61^. The stimulation then continued according to the default protocol (i.e., alternating 30 s phases of stimulation with biphasic impulse frequency of 25 Hz and 30 s pauses).

After a food-cue reactivity task (∼20 min; ^56, 60^), participants completed the EAT (∼40 min). The EAT ^16^, was adapted from ^62^ and assesses reward-related processes by the willingness to exert physical effort to gain food or money rewards depending on the difficulty and reward magnitude. Briefly, participants either worked for food or money tokens of either low (1 point) or high (10 points) magnitude. Difficulty also varied (easy vs. hard), leading to 8 possible trial combinations that were presented 6 times each. In each trial, participants saw a blue ball inside a vertical tube with a red horizontal line above it on the screen. The height of the red line indicated the level of difficulty and trial type (i.e., reward type and reward magnitude) was displayed in the upper right corner throughout the trial. To lift the blue ball above the red line, participants had to repeatedly press the right trigger button of an Xbox 360 controller (Microsoft Corporation, Redmond, WA); the faster the button was pressed, the higher the ball would go. For every second the ball was held above the line, money or food points were collected and the current score was displayed in the upper right corner of the screen. After each trial, participants were asked about how much they wanted the rewards and how much they exerted themselves via VAS. The task included 48 trials and two 15s breaks. During the EAT, taVNS was started in synchronization with the reward cue by the experimenter. Then, the stimulation continued with the default protocol while participants completed a reinforcement learning task (∼15 min; ^63^).

The task block was followed by VAS ratings. After removal of the taVNS electrode, the participants received their snack reward according to their achieved energy points and had time to eat as much as they liked during a short break. Then, participants answered the state VAS ratings for the last time. To complete the session, monetary winnings were paid out based on their earnings in the tasks. Both sessions followed the same standardized protocol and were conducted within an interval of 2-7 d (M = 4.68d ± 2.70). To evaluate the success of blinding, participants reported whether they received sham or taVNS at the end of each session. Their responses did not exceed the chance level (*recorded guesses*: 118, *correct guesses*: 63, *accuracy*: 53.3%, *p_binom_*=.52), suggesting successful blinding.

### Data analysis

#### Estimation of invigoration and maintenance of effort as a motivational index

To isolate invigoration and maintenance of effort as motivational indices, we segmented the behavioral data into work and rest segments (https://github.com/neuromadlab)^16^. We calculated taVNS effects using univariate mixed-effects models for our two dependent variables: effort invigoration and maintenance ^16^. Briefly, the models predicted each dependent variable based on the following dummy-coded variables: stimulation (taVNS, sham), reward type (food, money), reward magnitude (low, high), and difficulty (easy, hard), and the interaction between reward magnitude and difficulty. Additionally, we included interaction terms of stimulation with all other terms mentioned. To assess differential effects of taVNS in the MDD group compared to HCP, we included MDD diagnosis as a between-participant factor in interaction with the stimulation effect as well as reward magnitude and difficulty. Additionally, stimulation order (centered) was included as a nuisance variable. To account for deviations from fixed group effects, random slopes and intercepts were modeled for all predictors. Comparable models were used to investigate taVNS-induced changes in wanting.

To evaluate whether taVNS modulates the association between subjective ratings and motivation, we additionally used robust regression analysis that is preferable in the presence of heteroscedasticity and outliers ^64^ as is common in the rating data. Analogous to our previous work ^16^, we ran a robust regression (MATLAB robustfit, weight function huber) at the group level separately for the MDD and HCP groups as many participants had a restricted range in wanting ratings leading to uninformative individual slope estimates. Significance was assessed using permutation tests (with 10,000 iterations). We then compared the observed difference in slopes (taVNS – sham) to the null distribution to calculate *p*-values for the complete sample as well as HCP and MDD groups separately. Due to the advantages of robust regression, we used the same approach to compare increases in wanting and invigoration across trials and between taVNS and sham.

### Statistical threshold and software

For our analyses, we used a two-tailed α ≤.05 threshold. Mixed-effects analyses were conducted with lmerTest in R ^65^. Data was visualized using ggplot2 ^66^ and ggdist^67^. We processed data with MATLAB vR2021b and plotted results with R v4.1.1 (R Core Team, 2021).

## Results

To evaluate taVNS effects on effort invigoration and maintenance, we used mixed-effects models. First, we replicated the main task effects as participants worked more vigorously (invigoration: *b*= 6.2, *p*=.007) and harder for higher rewards; (maintenance: *b*= 9.3, *p*=.002). Participants also showed higher effort maintenance (*b*= 9.1, *p*=.0001), but not effort invigoration (*b*= -1.5, *p*=.35) for easy trials. In contrast to our previous study when participants were fasted, they now worked more vigorously (*b*= 5.1, *p*=.0005) and harder (*b*= 7.6, *p*<.001) for money as opposed to food. Regarding trial-wise subjective ratings, wanting was associated with both effort invigoration (*b*= 0.16, *p*<.001) and effort maintenance (*b*= 0.19, *p*<.001, Figure S1), whereas exertion was more strongly associated with effort maintenance (*b*= 0.35, *p*<.001) compared to invigoration (*b*= 0.08, *p*=.013), again replicating the pattern of our previous study.

Next, we compared performance between the HCP and MDD groups across conditions. To this end, we did not include interactions with group in this model. Participants with MDD did not exert less effort than HCP (Figure 2, all *p*s>.21) and performance was independent of symptom severity (as assessed by the BDI) and anhedonia (as assessed by the SHAPS (all *p*s>.38), see Tables S1-S4 for session specific differences). Adding sex, age, and BMI as covariates did not affect group- or stimulation-related inferences. In Session 1, patients with MDD reported lower wanting (*b*= -8.7, *p*=.032), but this difference was attenuated in Session 2 (*b*= -5.9, *p*=.33) and not significant across both sessions (*b*= -8.1, *p*=.078). Mirroring task performance, subjective ratings of exertion (S1: *b*= 0.9, *p*=.75, S2: *b*= 2.4, *p*=.41) were not different between groups. Again, neither symptom severity, nor anhedonia were specifically associated with wanting or exertion ratings (Table S5-S8).

**Figure 2.**
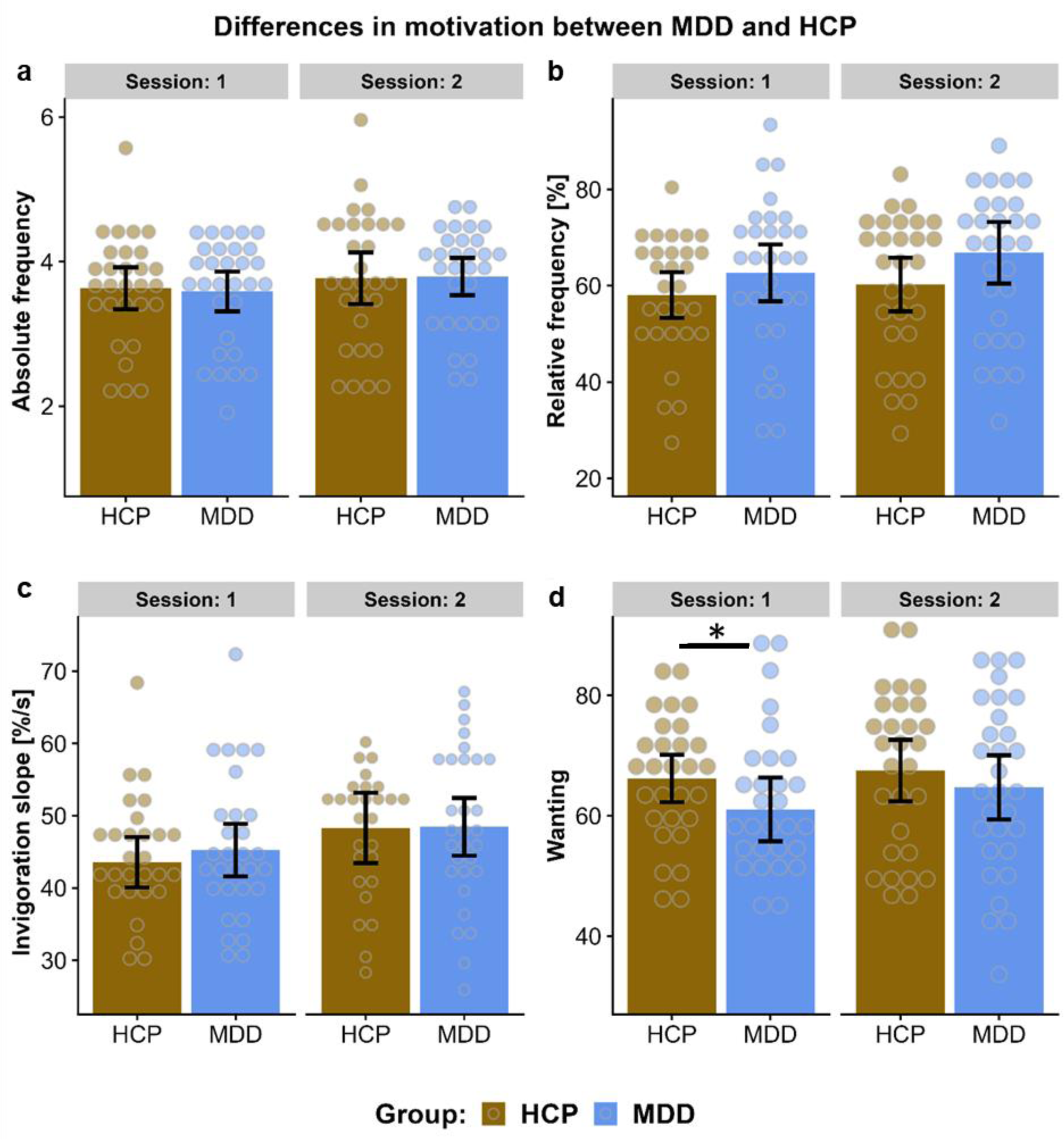
Patients with major depressive disorder (MDD) invest comparable effort for rewards but report lower wanting initially compared to healthy control participants (HCP). a: The absolute frequency of button presses showed no differences between groups in either session (Session 1: *b*= -0.05, *p*=.74, Session 2: *b*= -0.12, *p*=.46). b: Effort maintenance as assessed by the relative button press frequency throughout the trial was comparable across groups in Session 1 (*b*= 3.1, *p*=.33) and Session 2 (*b*= 4.5, *p*=.15). The nominally higher relative frequency was primarily due to the initial calibration at an individual level as absolute frequencies were nearly identical. c: There were no differences in invigoration slopes between groups in Session 1 (*b*= 1.9, *p*=.33) or Session 2 (*b*= 1.8, *p*=.59). d: Wanting was significantly lower in MDD compared to HCP in Session 1 (*b*= -7.0, *p*=.017), with a similar, non-significant trend in Session 2 (*b*= -2.4, *p*=.42).

### taVNS boosts the drive to work for less wanted rewards

We then sought to replicate our previous findings that taVNS increases effort invigoration in healthy participants and evaluated whether it generalizes to patients with MDD. In line with Neuser et al. ^16^, there was no taVNS-induced increase in effort maintenance (*b*= 1.1, *p*=.64, Table S10) across the sample. However, there was no taVNS-induced increase in invigoration (*b*= 0.02, *p*=.99, Table S9, Fig. 2a) across the sample and sessions. Nevertheless, in line with Neuser et al. ^16^, taVNS boosted invigoration (*b*= 5.6, *p*=.040, Cohen’s *d*_S1_= 0.55, Figure 2a) in Session 1, and invigoration did not change any further in Session 2 (Δinvigoration= 3.3, *p*=.12). Moreover, taVNS-induced changes were comparable in participants with vs. without MDD (Stim×Group: invigoration *p*=.87, maintenance *p*=. 80). Hence, participants who received taVNS in the first session continued to benefit from the stimulation but showed little additional change due to the acute stimulation in Session 2 (Figure 2a).

Next, we conducted a post hoc re-analysis of our previous data ^16^ to compare carryover effects. Although the previously reported taVNS-induced increase in invigoration was significant across both sessions, acute taVNS effects were also larger in the first session (Cohen’s *d*_S1_= 0.47, Cohen’s *d*_S2_= 0.01). Moreover, we replicated that taVNS increased the drive to work for less wanted rewards as indexed by a reduced correlation between wanting ratings and effort invigoration in the HCP group (HCP: *b*= -0.10, *p*_perm_=.0014, Fig. 3b-c). Notably, this taVNS-induced decrease in the slope was absent in patients with MDD (*b*= 0.05, *p*=.080). Whereas there was no difference in the correspondence of wanting and invigoration between groups in the sham condition (b= 0.05, *p*_perm_=.11), the HCP group had significantly lower correspondence between wanting and invigoration in the tVNS condition (*b*= -0.10, *p*=.008). To summarize, acute taVNS-induced increases in invigoration are replicably larger in Session 1, and these gains were more conserved and carried over to Session 2 in the present study.

**Figure 3.**
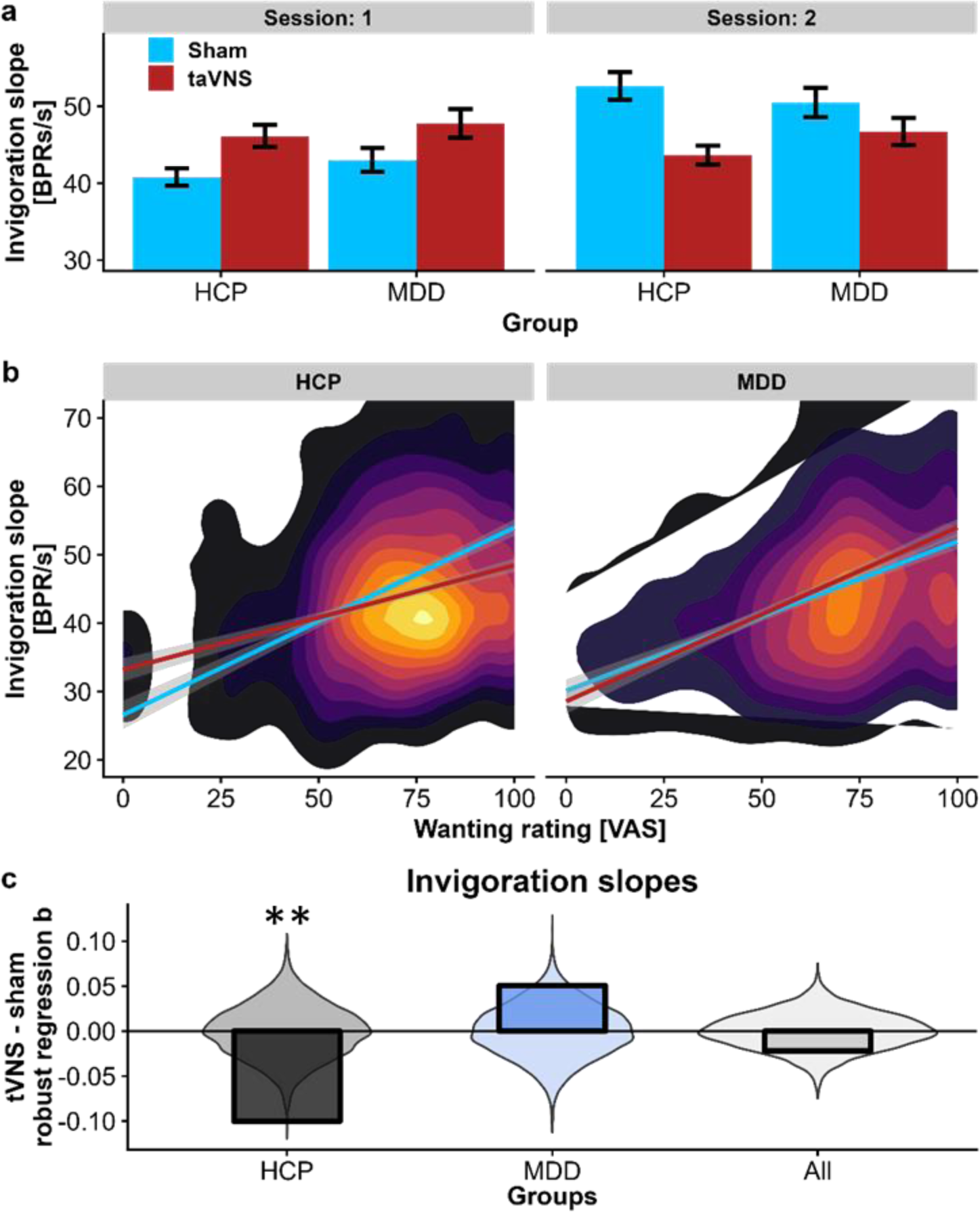
Vagus nerve stimulation (VNS) boosts the drive to work in Session 1 and for less wanted rewards in healthy control participants (HCP). a: Invigoration is increased in Session 1 compared to sham (*b*=5.6, *p*=.036) across both groups. In Session 2, invigoration did not change and differences from Session 1 were maintained (Δinvigoration= 3.3, *p*=.12). Participants receiving taVNS first are depicted in red in Session 1 and blue (sham) in Session 2 due to the crossover design of the study. Error bars depict 95% confidence intervals at the trial level. b: During taVNS, effort invigoration becomes less dependent on wanting in HCP (*p*_perm_=.0014), but not in patients with MDD (where the slope tends to increase, *p*_perm_=.089). Wanting ratings are shown as 2d-density polygon, where brighter colors indicate a higher density of data. c: Distributions of the permutated associations between invigoration and wanting during taVNS vs. sham. We fitted robust regression coefficients, *b*, after permuting the labels for taVNS vs. sham stimulation, and calculated the difference in slopes (colored bars: dark gray = HCP, light blue = MDD, light gray = combined sample) to a permuted null distribution (violin plots in the background in gray). This permutation test showed a significant main effect of taVNS in HCP, but not patients with MDD. BPR/s = button press rate in % per s.

### taVNS durably enhances motivation and gains extend to wanting in depression

To better understand how taVNS-induced increases in Session 1 are translated to lasting motivational differences in Session 2, we further explored trial-based dynamics of taVNS effects. We reasoned that stimulation-induced changes in behavior and ratings across trials might track behavioral adaptations and that taVNS-induced gains might be linked to durable changes in the subjective value of the rewards at stake. To investigate trial-based dynamics, we estimated the effect of taVNS (vs. sham) on changes in effort invigoration and wanting within sessions (i.e., by estimating trial slopes capturing changes over trials). In line with an instrumental conditioning effect, taVNS induced stronger increases in effort invigoration (*p*_perm_=.005) and wanting (*p*_perm_=.011) during the first session in patients with MDD, but not HCP (invigoration: *p*_perm_=.96; wanting: *p*_perm_=.90). When including continuous symptom severity instead of group, participants with a higher BDI showed lower trial-wise increases in invigoration (*r*=.37, *p*=.046, Figure S1-2) during Session 1 (with a comparable trend for wanting, *r*=.28, p=.14). During taVNS, this association of increases in invigoration with BDI was fully attenuated (*r*=.00, *p*=.99), indicating that taVNS normalized motivation in Session 1 (for associations with other baseline characteristics, see Figure S1). Notably, invigoration and wanting plateaued at the end of Session 1 and there were no incremental changes in Session 2 (invigoration: *b*= 3.3, *p*=.12, wanting: *b* = -0.29, *p*=. 88), suggesting that gains were largely preserved across sessions and only marginally influenced by acute stimulation after learning on the task. Hence, we also assessed whether taVNS-induced changes between sessions were affected by the delay. While we observed nominally lower indices when sessions were further apart, the association was not significant (invigoration: *b*= -0.97, *t*(55)= -1.48, *p*=.15; wanting: *b*= -0.97, *t*(55)= -1.37, *p*=.17, Figure S3) and a reanalysis of the previous HCP data did not show the same pattern (invigoration: *b*= 1.1, *t*(79)= 1.51, *p*=.13; wanting: *b*= -0.81, *t*(79)= -1.28, *p*=.21). Taken together, our results suggest that taVNS induces rapid gains in invigoration and wanting in patients with MDD that persist into Session 2.

**Figure 3.**
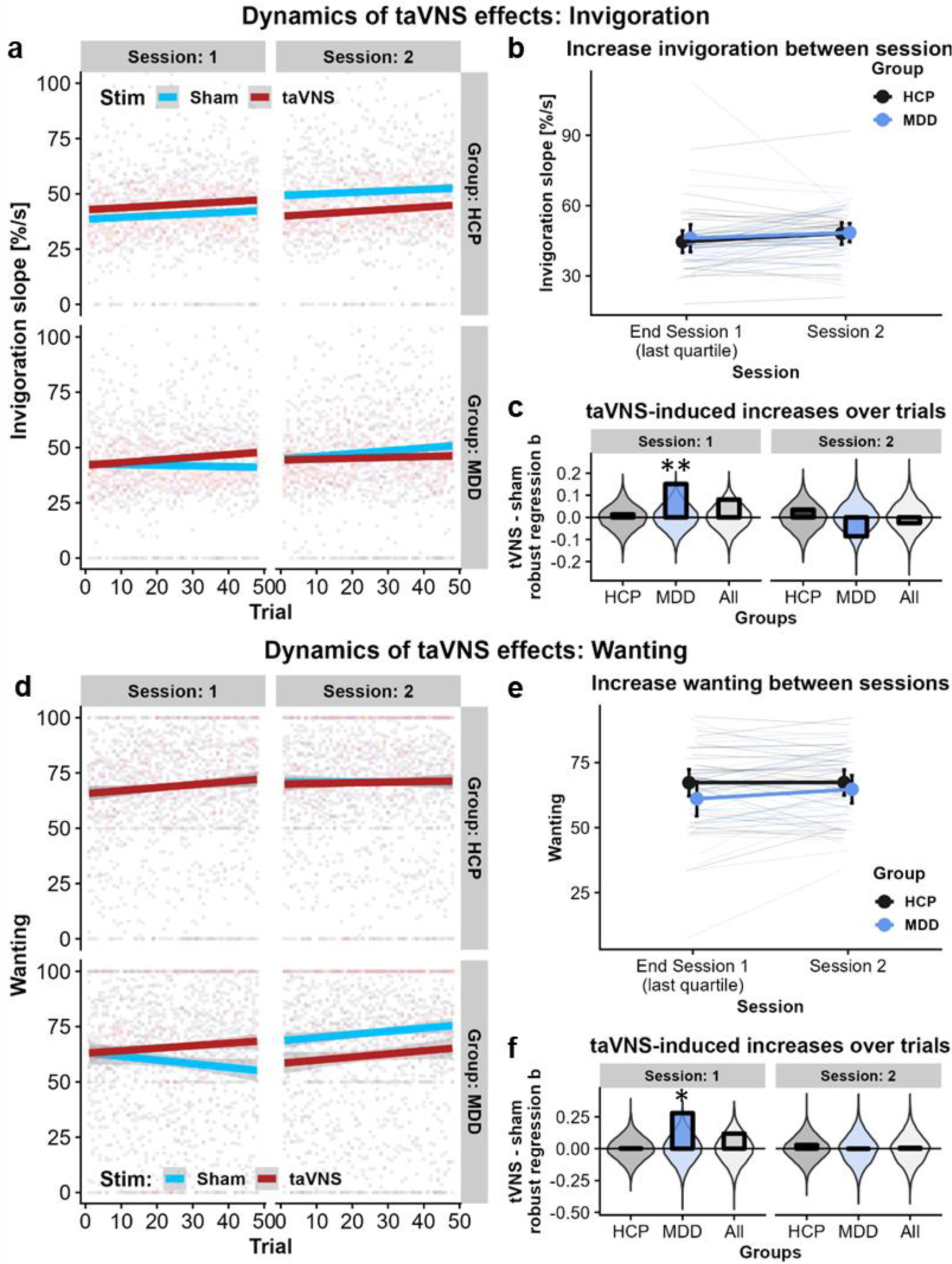
Transcutaneous auricular vagus nerve stimulation (taVNS) increases effort invigoration and wanting across trials in patients with major depressive disorder (MDD). a: Trajectories of effort invigoration across trials split by stimulation, session, and group. In Session 1, patients with MDD show stronger taVNS-induced increases in invigoration across trials (*p*_perm_=.005), indicating instrumental learning. b: Across participants, the conditioned increase in invigoration in Session 1 does not change in Session 2 (*p*=.12). c: taVNS-induced increases in invigoration across trials are observed in patients with MDD, but not HCP compared to permuted null distributions (colored bars: dark gray = HCP, light blue = MDD, light gray = combined sample). d: Trajectories of wanting ratings across trials split by stimulation, session, and group. In Session 1, patients with MDD show stronger taVNS-induced increases across trials (*p*_perm_=.011) indicating instrumental conditioning. e: Learned evaluation of the reward at stake as indexed by wanting in the last quarter of the experiment in Session 1 does not change in Session 2 (*p*=.88) across participants. f: taVNS-induced increases in invigoration slope across trials are observed in patients with MDD, but not HCP compared to permuted null distributions.

## Discussion

Loss of motivation is a pervasive symptom of MDD that is often not effectively treated by the current first line of treatment. To this end, we evaluated acute effects of taVNS on the willingness to work for rewards in participants with and without MDD. First, we replicated that taVNS boosts invigoration in both groups during the first session, corroborating our previously reported findings in healthy participants ^16^. Second, we did not replicate that acute taVNS leads to an increase in invigoration across both sessions. Instead, taVNS-induced gains in effort invigoration persisted into the second session, demonstrating durable increases that could reflect instrumental conditioning. In support of this interpretation, we observed increases in invigoration and wanting in patients with MDD across trials in the first session, indicating that taVNS may facilitate effort-related learning processes that translate to the subjective evaluation. Surprisingly, these transfer effects were stable and largely resistant to the effects of acute stimulation during the second session. Third, we also replicated an increased invigoration for less wanted rewards in HCP, but not in patients with MDD. This discrepancy corroborates differences in the dynamics of learning during the effort task, where taVNS facilitated a rapid improvement of motivation during the first session that translated immediately to changes in subjective wanting in patients with MDD, but not in HCP. Taken together, our study demonstrates that acute taVNS-induced changes in invigoration and wanting may facilitate value-related learning processes that could be beneficial for the development of novel treatments for motivational symptoms of MDD.

In line with our previous results in healthy participants, taVNS increased invigoration in both groups in the first session. However, taVNS-induced increases in invigoration persisted into the second session and were no longer substantially altered by the acute stimulation, indicating longer-lasting learning effects. We then reanalyzed our previously collected data ^16^ and observed larger taVNS-induced effects in the first session as well that were comparable to the new sample. In contrast, carryover effects were more pronounced compared to Neuser et al.^16^. These differences might be due to changes in our procedure. First, more participants completed their second session two days after the first, leading to a shorter time interval between sessions. Second, to ease the recruitment of patients with MDD during the pandemic, participants completed the sessions in a non-fasting state corresponding to neither hungry nor full^68^ and at different times of day ^69^. Persistent effects of taVNS due to learning are in line with rodent studies showing that VNS improves memory persistence ^70^. In humans, taVNS-enhanced memories were primarily reported for emotional stimuli and episodic content ^46, 47, 71, 72^. Likewise, taVNS has been shown to reduce learning rates which may also reflect longer-lasting memory traces for rewards ^63^. The interpretation that taVNS leads to a differentially learned memory trace is strengthened by the observation that taVNS-induced gradual increases of invigoration and wanting in patients with MDD in the first session. Crucially, the transfer of a heightened reward drive with corresponding changes in the subjective wanting of rewards in patients with MDD is highly promising for potential interventions. Such trial-to-trial increases in effort invigoration might be explained by enhanced motor learning which has been observed in rats ^73, 74^ and translated for clinical use in the recovery of motor function after a stroke ^75^. In healthy humans, the evidence for taVNS effects on reinforcement learning is still mixed ^63, 76^, but a small study in patients with epilepsy also reported taVNS-induced improvements in learning across trials ^77^. Taken together, taVNS might lead to changes in effort-related learning processes, and harnessing these effects might help normalize motivation in patients with MDD to provide a lasting boost in reward-related behavior.

In support of a largely subconscious boost of motivational drive, we replicated that taVNS enhances invigoration for less wanted rewards in healthy participants ^16^. Hence, taVNS might enhance the utility to work for rewards regardless of the expected benefit which would be in line with changes in monoaminergic signaling ^78^. Intriguingly, this reduction in the utility slope was only observed in healthy participants, and not in patients with MDD. To better understand this discrepancy, we conducted additional analyses within Session 1, demonstrating trial-wise increases in wanting during taVNS in patients with MDD. This gradual change in subjective ratings that we did not observe in healthy participants might reflect a normalization of reward-related dysfunctions in patients with MDD ^21, 25, 32, 79^. Previous work in patients with MDD has shown that their behavior is less sensitive to manipulations of reward values ^80^ and taVNS may rapidly improve sensitivity to task-related performance feedback as a mechanism of learning. Likewise, taVNS may enhance sensitivity to interoceptive signals ^12, 17^ contributing to effortful behavior and the subjective evaluation of costs and benefits of action ^16, 81, 82^. Therefore, alterations in interoception that have been recently linked to depression ^83–86^ might provide a mechanism for taVNS to rapidly improve instrumental learning, both in terms of invigoration and subjective wanting. Since MDD is also characterized by dysregulation of the autonomic nervous system ^87, 88^ and the monoaminergic system^35, 89–91^, taVNS may help normalize signaling in patients with MDD. Consequently, taVNS might reveal differential modes of action in healthy populations compared to patients with MDD which may ultimately contribute to an improved understanding of the role of vagal afferent signals in regulating motivation in both health and disease.

To guide future research, several limitations of the current study must be considered. First, although persistent taVNS-induced gains in motivation are desirable for interventions, our crossover design was not optimized to resolve such carryover effects. Our post hoc analyses indicate that longer intervals between sessions may attenuate carryover effects (Figure S2). However, this pattern was not visible in our previous data ^16^. Therefore, future work should investigate carryover effects with more longitudinal sessions, including the pressing clinical questions how long taVNS-induced changes in invigoration and wanting last and how long-term changes in motivation could be facilitated. Second, to improve accessibility for patients with MDD, experimental sessions took place throughout the day, not only after an overnight fast as in our previous studies ^16, 54, 60, 63^. Hence, metabolic states were more variable among participants (i.e., sessions took place 3 to 5 h after the last meal). Considering the role of vagal signaling in conferring the current state of the body ^11^, the effects of taVNS might conceivably depend on the metabolic state ^68^. Third, patients with MDD did not exert less effort compared to HCP as previous research has suggested ^18, 20^. Here, we predominantly recruited patients with mild to medium severity of depression (∼71%) who were receiving treatment. MDD is a heterogeneous disorder with distinct subgroups and symptom profiles ^26^, and impaired motivation is one facet of anhedonia ^18, 20, 32^. Nevertheless, patients with MDD reported lower wanting of rewards during the first session. Larger samples including patients with greater symptom severity and, ideally, without concurrent medication with SSRIs might yield more robust associations with behavior, even though this would reduce the representative of the patient sample. Fourth, although including the current medication (i.e., taking antidepressants vs. no medication) did not alter the reported results, taVNS might interact differently with antidepressants depending on whether they primarily target the dopamine or serotonin system ^30^. Since the current study is not sufficiently powered to evaluate differences between antidepressant classes, future studies are needed to investigate potential interactions of taVNS with commonly prescribed antidepressants.

Motivational symptoms of MDD are difficult to treat and we tested whether previously discovered acute invigorating effects of taVNS also occur in patients with MDD. Accordingly, we found that taVNS boosted invigoration across groups in the first session and taVNS-induced gains even persisted into the second session. In patients with MDD, taVNS-induced gains in invigoration evolved over trials and were mirrored in increases in wanting, suggesting an instrumental conditioning of subjective value that also persisted into the second session. Notably, we also replicated that taVNS boosted invigoration for less wanted rewards in healthy participants while patients with MDD showed instant improvements in subjective ratings of reward. To conclude, our results highlight distinct dynamics of instrumental learning elicited by taVNS in HCP and MDD. Specifically, taVNS-induced instrumental conditioning of effort, leading to durable effects on invigoration and wanting, appears highly promising for future motivational treatments, where taVNS could be used as an adjuvant in behavioral modules of therapy.

## Data Availability

The datasets used during the current study are available from the corresponding author on reasonable request.

## Acknowledgement

We thank Franziska Kräutlein and Larissa Katz for help with data acquisition as well as Wiebke Ringels for support in processing the data. The study was supported by the University of Tübingen, Faculty of Medicine fortune grant 2453-0-0, Daimler & Benz Foundation 32-04/19, and the Deutsche Forschungsgemeinschaft (DFG), grants KR 4555/7-1, KR 4555/9-1, & KR 4555/10-1.

## Author contributions

NBK was responsible for the study concept and design. MF, JK, & WML recruited participants and collected data under supervision by NBK. NBK conceived the method and MF & AK processed the data. MF, AK, & NBK performed the data analysis. MF, AK, & NBK wrote the manuscript. All authors contributed to the interpretation of findings, provided critical revision of the manuscript for important intellectual content, and approved the final version for publication.

