## Supplementary Information for "Non-invasive vagus nerve stimulation conditions increased invigoration and wanting in depression"

Venusberg Campus 1, 53127 Bonn, Germany

### Figures

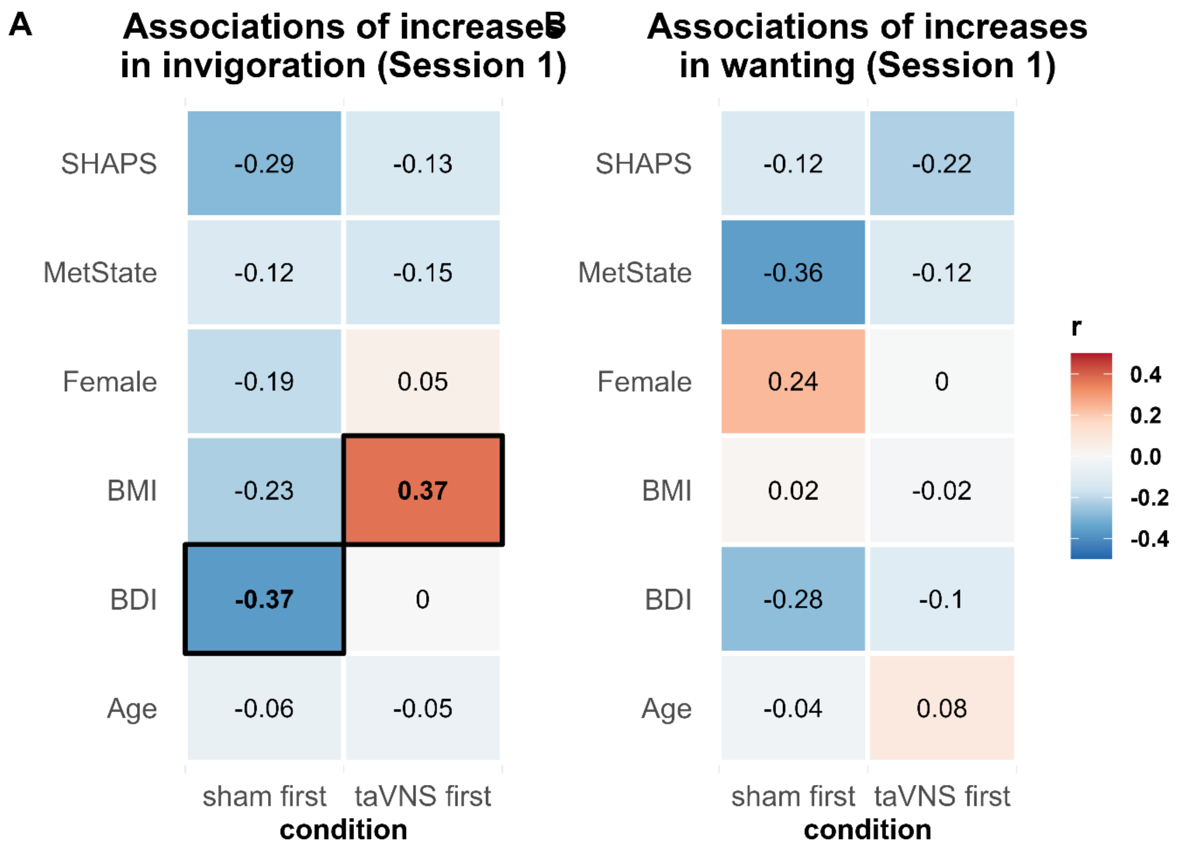


**Figure S1:** taVNS-induced trial-wise increases in invigoration during session 1 depend on BMI and reduce differences related to depressive symptoms. A) Correlations of trial-wise increased in invigoration during session 1 with baseline characteristics separated for sham and taNVS. In the sham condition, more depressive symptoms are associated with decreased in invigoration from trial-to-trial ($r(28)= .37$, $p= .046$). In contrast, during taVNS symptom severity does not affect changes across trials ($r(27)= .001$, $p= .99$). Moreover, taVNS induced increased are higher in participants with an increased BMI ($r(27)= .37$, $p= .046$). SHAPS = Snaith Hamilton Pleasure Scale, BMI = Body mass index, BDI = Beck Depression Inventory, taVNS = transcutaneous vagus nerve stimulation.

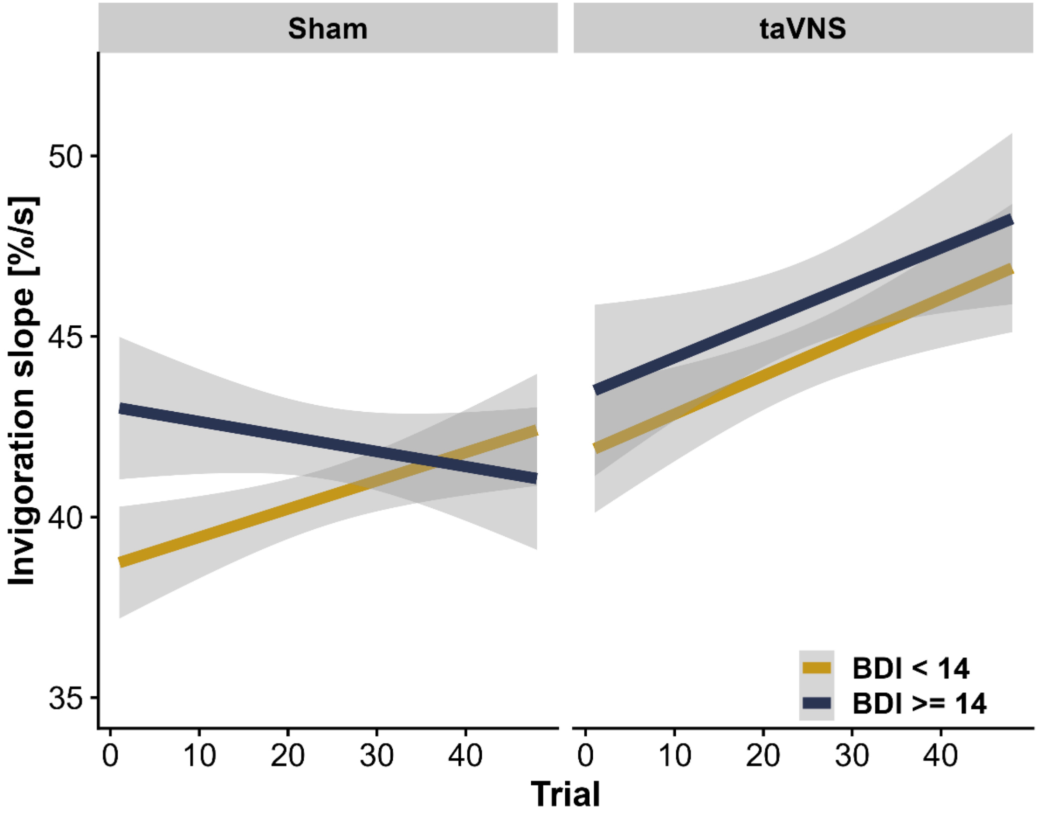


**Figure S2:** In Session 1, trial-wise increases in invigoration slope depend on depressive symptom severity (Beck Depression Inventory, BDI) in the sham condition ($r(28)= .37$, $p= .046$), but not the transcutaneous auricular vagus nerve stimulation (taVNS) condition ($r(27)= .001$, $p= .99$).

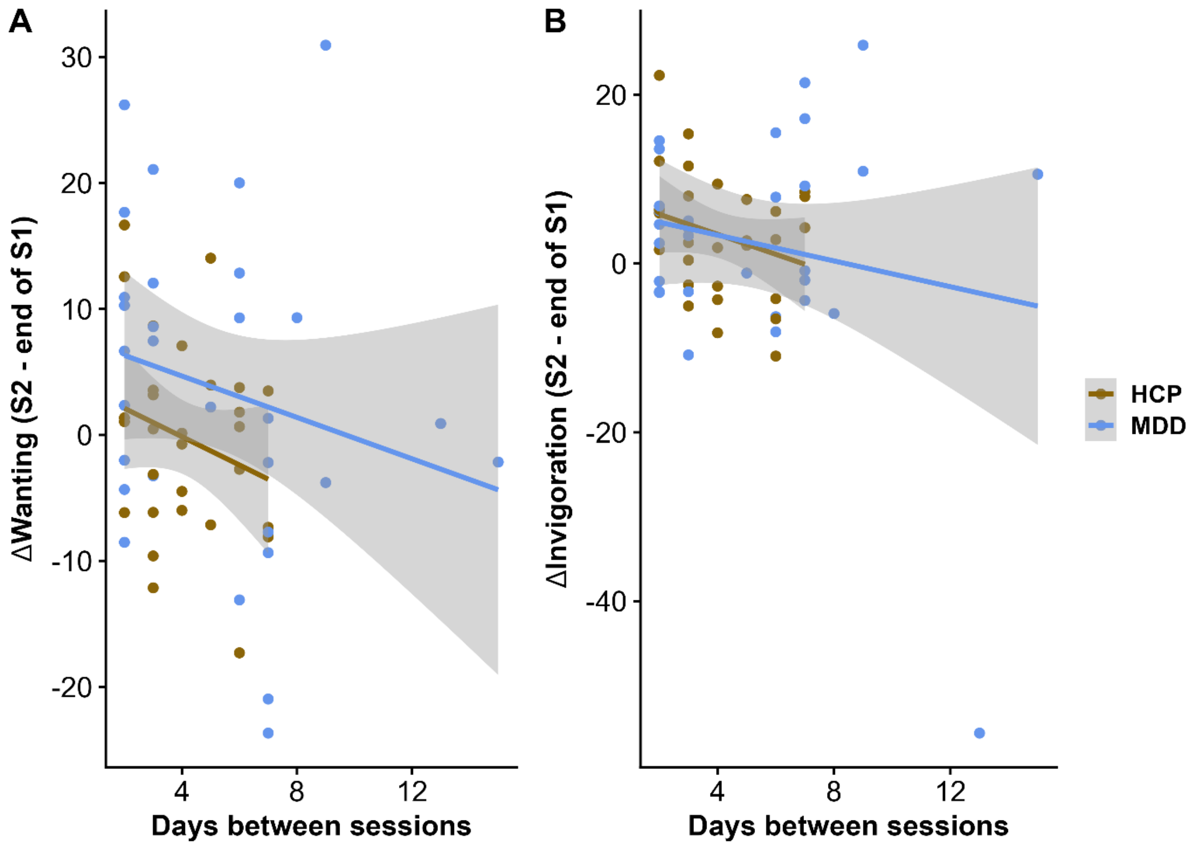


**Figure S3:** Carry over effects between session for wanting and invigoration depend on the number of days between session with stronger losses if sessions are further apart. A) Differences in wanting between the end of session 1 and session 2 are numerically more negative (i.e., any gains during session 1 are lost again) the further apart the sessions were completed (*b*= -2.6, *t*(55)= -1.48, *p*= 0.15). B) Differences in invigoration between the end of session 1 and session 2 are numerically more negative (i.e., any gains during session 1 are lost again) the further apart the sessions were completed (*b*= -2.6, *t*(55)= -1.37, *p*= 0.17).

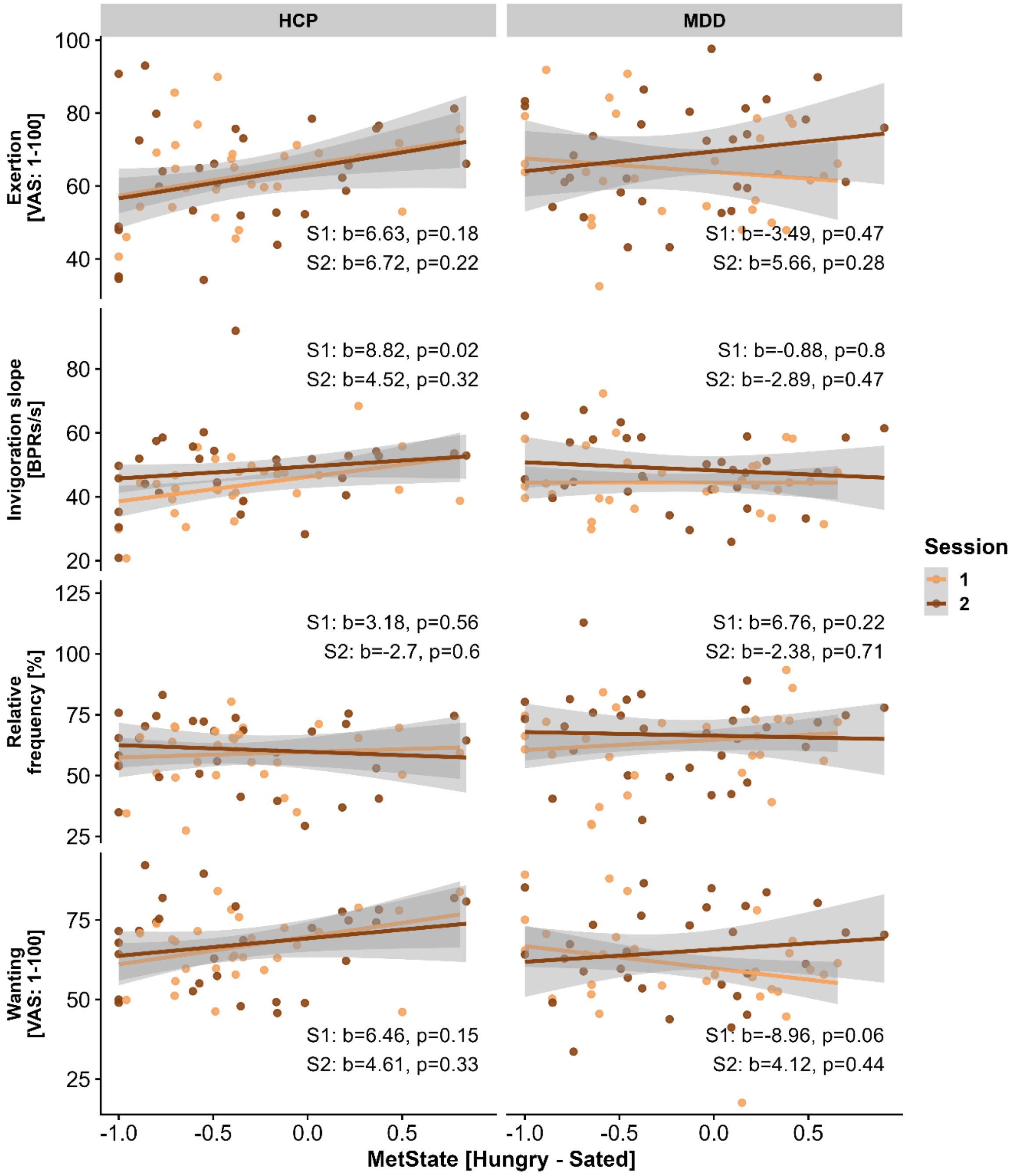


**Figure S4:** In healthy participants, motivation depends on the current metabolic state (Hungry – Sated) in session 1.

**Tables**

| | Invigoration Slope | | | Invigoration Slope | | | Invigoration Slope | | |
|---|---|---|---|---|---|---|---|---|---|
| *Predictors* | *Estimates* | *p* | *df* | *Estimates* | *p* | *df* | *Estimates* | *p* | *df* |
| (Intercept) | 34.6 | **<0.001** | 49.0 | 35.5 | **<0.001** | 37.4 | 35.6 | **<0.001** | 37.1 |
| Stim [taVNS] | 5.6 | **0.044** | 56.7 | 5.7 | **0.042** | 56.3 | 5.6 | **0.046** | 55.1 |
| RewMag [High] | 8.4 | **0.003** | 29.3 | 8.4 | **0.003** | 29.4 | 8.4 | **0.003** | 29.3 |
| Diff [Hard] | 0.5 | 0.728 | 39.1 | 0.5 | 0.728 | 39.2 | 0.5 | 0.727 | 39.1 |
| Money [Money] | 4.7 | **0.001** | 58.0 | 4.7 | **0.001** | 58.0 | 4.7 | **0.001** | 58.0 |
| Stim [taVNS] × RewMag [High] | -2.4 | 0.400 | 46.6 | -2.4 | 0.400 | 46.7 | -2.4 | 0.400 | 46.7 |
| Stim [taVNS] × Diff [Hard] | 1.0 | 0.648 | 94.4 | 1.0 | 0.648 | 94.2 | 1.0 | 0.644 | 92.8 |
| RewMag [High] × Diff [Hard] | -1.5 | 0.541 | 29.2 | -1.5 | 0.541 | 29.1 | -1.5 | 0.541 | 29.3 |
| Stim [taVNS] × RewMag [High] × Diff [Hard] | -0.6 | 0.853 | 65.4 | -0.6 | 0.853 | 65.2 | -0.6 | 0.853 | 62.6 |
| Group [MDD] | 1.9 | 0.327 | 47.5 | | | | | | |
| BDI | | | | 1.5 | 0.111 | 44.2 | | | |
| SHAPS | | | | | | | 2.1 | **0.027** | 47.0 |

**Table S3**: Invigoration slope does not differ between participants with major depressive disorder (MDD) and healthy control participants (HCP) in session 1. Results from a linear mixed effects model. BDI = Beck’s Depression Inventory. SHAPS = Snaith Hamilton pleasure scale

| | Invigoration Slope | | | Invigoration Slope | | | Invigoration Slope | | |
|---|---|---|---|---|---|---|---|---|---|
| *Predictors* | *Estimates* | *p* | *df* | *Estimates* | *p* | *df* | *Estimates* | *p* | *df* |
| (Intercept) | 43.2 | **<0.001** | 41.4 | 44.3 | **<0.001** | 31.5 | 44.2 | **<0.001** | 31.3 |
| Stim [taVNS] | -6.2 | 0.067 | 54.6 | -6.5 | 0.060 | 53.7 | -6.2 | 0.067 | 55.2 |
| RewMag [High] | 7.7 | **<0.001** | 29.5 | 7.7 | **<0.001** | 29.3 | 7.7 | **<0.001** | 29.8 |
| Diff [Hard] | 1.7 | 0.328 | 46.9 | 1.7 | 0.327 | 46.5 | 1.7 | 0.324 | 46.7 |
| Money [Money] | 5.8 | **<0.001** | 58.1 | 5.8 | **<0.001** | 58.1 | 5.8 | **<0.001** | 58.1 |
| Stim [taVNS] × RewMag [High] | 1.3 | 0.713 | 50.5 | 1.3 | 0.713 | 50.4 | 1.3 | 0.713 | 50.6 |
| Stim [taVNS] × Diff [Hard] | -2.4 | 0.295 | 107.5 | -2.4 | 0.294 | 107.1 | -2.4 | 0.293 | 108.0 |
| RewMag [High] × Diff [Hard] | -0.7 | 0.747 | 30.9 | -0.7 | 0.746 | 30.1 | -0.7 | 0.747 | 31.9 |
| Stim [taVNS] × RewMag [High] × Diff [Hard] | 1.7 | 0.583 | 80.3 | 1.7 | 0.584 | 79.2 | 1.7 | 0.583 | 82.0 |
| Group [MDD] | 1.8 | 0.487 | 52.1 | | | | | | |
| BDI | | | | 2.3 | 0.080 | 48.2 | | | |
| SHAPS | | | | | | | 1.5 | 0.272 | 52.1 |

**Table S4:** Invigoration slope does not differ between participants with major depressive disorder (MDD) and healthy control participants (HCP) in session 2. Results from a linear mixed effects model**.** BDI = Beck's Depression Inventory. SHAPS = Snaith Hamilton pleasure scale.

| | Effort maintenance | | | Effort maintenance | | | Effort maintenance | | |
|---|---|---|---|---|---|---|---|---|---|
| *Predictors* | *Estimates* | *p* | *df* | *Estimates* | *p* | *df* | *Estimates* | *p* | *df* |
| (Intercept) | 52.6 | **<0.001** | 49.7 | 54.1 | **<0.001** | 38.3 | 54.2 | **<0.001** | 38.3 |
| Stim [taVNS] | 3.0 | 0.469 | 56.5 | 3.1 | 0.453 | 56.2 | 2.8 | 0.495 | 56.2 |
| RewMag [High] | 11.8 | **0.001** | 30.5 | 11.8 | **0.001** | 30.5 | 11.8 | **0.001** | 30.5 |
| Diff [Hard] | -10.3 | **<0.001** | 29.2 | -10.3 | **<0.001** | 29.3 | -10.3 | **<0.001** | 29.2 |
| Money [Money] | 7.7 | **<0.001** | 58.0 | 7.7 | **<0.001** | 58.0 | 7.7 | **<0.001** | 58.0 |
| Stim [taVNS] × RewMag [High] | -4.2 | 0.217 | 43.6 | -4.2 | 0.217 | 43.5 | -4.2 | 0.217 | 43.5 |
| Stim [taVNS] × Diff [Hard] | 1.4 | 0.675 | 55.5 | 1.4 | 0.675 | 55.5 | 1.4 | 0.675 | 55.5 |
| RewMag [High] × Diff [Hard] | 2.8 | **0.032** | 91.8 | 2.8 | **0.033** | 87.2 | 2.8 | **0.031** | 94.2 |
| Stim [taVNS] × RewMag [High] × Diff [Hard] | 1.8 | 0.435 | 53.2 | 1.8 | 0.436 | 53.3 | 1.8 | 0.435 | 53.1 |
| Group [MDD] | 3.1 | 0.326 | 51.8 | | | | | | |
| BDI | | | | 2.0 | 0.207 | 48.7 | | | |
| SHAPS | | | | | | | 0.1 | 0.951 | 53.9 |

**Table S7**: Effort maintenance (RelEffort) does not differ between participants with major depressive disorder (MDD) compared to healthy control participants (HCP) in session 1. Results from a linear mixed-effects model**.** BDI = Beck's Depression Inventory. SHAPS = Snaith Hamilton pleasure scale

| | Effort maintenance | | | Effort maintenance | | | Effort maintenance | | |
|---|---|---|---|---|---|---|---|---|---|
| *Predictors* | *Estimates* | *p* | *df* | *Estimates* | *p* | *df* | *Estimates* | *p* | *df* |
| (Intercept) | 56.2 | **<0.001** | 39.8 | 58.6 | **<0.001** | 32.7 | 58.4 | **<0.001** | 32.6 |
| Stim [taVNS] | -1.1 | 0.820 | 56.2 | -1.3 | 0.783 | 56.0 | -0.9 | 0.853 | 55.8 |
| RewMag [High] | 9.1 | **0.001** | 28.0 | 9.1 | **0.001** | 28.0 | 9.1 | **0.001** | 28.1 |
| Diff [Hard] | -6.9 | **0.006** | 28.5 | -6.9 | **0.006** | 28.5 | -6.9 | **0.006** | 28.5 |
| Money [Money] | 7.9 | **<0.001** | 58.0 | 7.9 | **<0.001** | 58.0 | 7.9 | **<0.001** | 58.0 |
| Stim [taVNS] × RewMag [High] | 3.1 | 0.449 | 54.7 | 3.1 | 0.449 | 54.7 | 3.1 | 0.449 | 54.7 |
| Stim [taVNS] × Diff [Hard] | -5.8 | 0.095 | 56.7 | -5.8 | 0.095 | 56.7 | -5.8 | 0.095 | 56.7 |
| RewMag [High] × Diff [Hard] | 4.0 | **0.029** | 28.4 | 4.0 | **0.029** | 28.4 | 4.0 | **0.029** | 28.4 |
| Stim [taVNS] × RewMag [High] × Diff [Hard] | 1.2 | 0.595 | 54.5 | 1.2 | 0.595 | 54.5 | 1.2 | 0.595 | 54.5 |
| Group [MDD] | 4.5 | 0.145 | 44.5 | | | | | | |
| BDI | | | | 3.1 | **0.039** | 41.6 | | | |
| SHAPS | | | | | | | 1.3 | 0.398 | 45.4 |

**Table S8**: Effort maintenance (RelEffort) does not differ between participants with major depressive disorder (MDD) compared to healthy control participants (HCP) in session 2. Results from a linear mixed-effects model**.** BDI = Beck’s Depression Inventory. SHAPS = Snaith Hamilton pleasure scale

| | Wanting | | | Wanting | | | Wanting | | |
|---|---|---|---|---|---|---|---|---|---|
| *Predictors* | *Estimates* | *p* | *df* | *Estimates* | *p* | *df* | *Estimates* | *p* | *df* |
| (Intercept) | 50.8 | **<0.001** | 44.1 | 47.1 | **<0.001** | 34.3 | 47.0 | **<0.001** | 34.2 |
| Stim [taVNS] | 3.9 | 0.358 | 51.5 | 4.1 | 0.344 | 50.8 | 4.2 | 0.330 | 52.2 |
| RewMag [High] | 22.0 | **<0.001** | 31.6 | 22.0 | **<0.001** | 31.6 | 22.0 | **<0.001** | 31.6 |
| Diff [Hard] | -10.1 | **<0.001** | 28.9 | -10.1 | **<0.001** | 29.1 | -10.1 | **<0.001** | 29.0 |
| Money [Money] | 15.9 | **<0.001** | 58.0 | 15.9 | **<0.001** | 58.0 | 15.9 | **<0.001** | 58.0 |
| Stim [taVNS] × RewMag [High] | -5.9 | 0.172 | 47.8 | -5.9 | 0.172 | 47.8 | -5.9 | 0.172 | 47.8 |
| Stim [taVNS] × Diff [Hard] | 3.6 | 0.256 | 51.0 | 3.6 | 0.255 | 51.2 | 3.6 | 0.256 | 51.1 |
| RewMag [High] × Diff [Hard] | 4.6 | **0.048** | 29.2 | 4.6 | **0.048** | 29.3 | 4.6 | **0.048** | 29.2 |
| Stim [taVNS] × RewMag [High] × Diff [Hard] | 0.5 | 0.863 | 61.4 | 0.5 | 0.863 | 61.7 | 0.5 | 0.863 | 61.5 |
| Group [MDD] | -7.1 | **0.017** | 55.4 | | | | | | |
| BDI | | | | -1.3 | 0.402 | 55.4 | | | |
| SHAPS | | | | | | | -2.1 | 0.174 | 56.0 |

**Table S5**: Wanting is lower in participants with major depressive disorder (MDD) compared to healthy control participants (HCP) in session 1. Results from a linear mixed effects model**.** BDI = Beck's Depression Inventory. SHAPS = Snaith Hamilton pleasure scale

| | Wanting | | | Wanting | | | Wanting | | |
|---|---|---|---|---|---|---|---|---|---|
| *Predictors* | *Estimates* | *p* | *df* | *Estimates* | *p* | *df* | *Estimates* | *p* | *df* |
| (Intercept) | 53.8 | **<0.001** | 43.6 | 52.7 | **<0.001** | 35.1 | 52.6 | **<0.001** | 35.1 |
| Stim [taVNS] | -6.2 | 0.182 | 54.3 | -6.5 | 0.171 | 55.2 | -6.4 | 0.175 | 55.0 |
| RewMag [High] | 20.1 | **<0.001** | 28.9 | 20.1 | **<0.001** | 28.9 | 20.1 | **<0.001** | 28.9 |
| Diff [Hard] | -5.0 | **0.001** | 29.0 | -5.0 | **0.001** | 28.9 | -5.0 | **0.001** | 29.0 |
| Money [Money] | 14.7 | **<0.001** | 58.0 | 14.7 | **<0.001** | 58.0 | 14.7 | **<0.001** | 58.0 |
| Stim [taVNS] × RewMag [High] | 6.9 | 0.116 | 56.0 | 6.8 | 0.116 | 56.1 | 6.9 | 0.116 | 56.0 |
| Stim [taVNS] × Diff [Hard] | -3.4 | 0.128 | 57.9 | -3.4 | 0.128 | 57.9 | -3.4 | 0.128 | 57.9 |
| RewMag [High] × Diff [Hard] | 2.6 | 0.116 | 56.2 | 2.6 | 0.117 | 55.7 | 2.6 | 0.116 | 56.0 |
| Stim [taVNS] × RewMag [High] × Diff [Hard] | 1.4 | 0.530 | 116.7 | 1.4 | 0.530 | 116.5 | 1.4 | 0.530 | 116.6 |
| Group [MDD] | -2.4 | 0.415 | 55.5 | | | | | | |
| BDI | | | | 0.4 | 0.801 | 55.4 | | | |
| SHAPS | | | | | | | -0.9 | 0.543 | 56.4 |

**Table S6**: Wanting does not differ between participants with major depressive disorder (MDD) compared to healthy control participants (HCP) in session 2. Results from a linear mixed effects model**.** BDI = Beck's Depression Inventory. SHAPS = Snaith Hamilton pleasure scale

| | Exertion [Group] | | | Exertion [BDI] | | | Exertion [SHAPS] | | |
|---|---|---|---|---|---|---|---|---|---|
| *Predictors* | *Estimates* | *p* | *df* | *Estimates* | *p* | *df* | *Estimates* | *p* | *df* |
| (Intercept) | 54.6 | **<0.001** | 42.5 | 55.1 | **<0.001** | 34.6 | 55.0 | **<0.001** | 34.2 |
| Stim [taVNS] | 0.2 | 0.961 | 45.5 | 0.2 | 0.970 | 45.5 | 0.2 | 0.956 | 44.5 |
| RewMag [High] | 17.8 | **<0.001** | 29.2 | 17.8 | **<0.001** | 29.2 | 17.8 | **<0.001** | 29.1 |
| Diff [Hard] | -12.1 | **<0.001** | 29.3 | -12.1 | **<0.001** | 29.3 | -12.1 | **<0.001** | 29.3 |
| Money [Money] | 10.7 | **<0.001** | 58.0 | 10.7 | **<0.001** | 58.0 | 10.7 | **<0.001** | 57.9 |
| Stim [taVNS] × RewMag [High] | -6.2 | 0.175 | 45.2 | -6.2 | 0.175 | 45.2 | -6.2 | 0.175 | 45.1 |
| Stim [taVNS] × Diff [Hard] | 1.9 | 0.628 | 56.4 | 1.9 | 0.628 | 56.4 | 1.9 | 0.628 | 56.4 |
| RewMag [High] × Diff [Hard] | 5.7 | 0.059 | 29.1 | 5.7 | 0.059 | 29.1 | 5.7 | 0.059 | 29.1 |
| Stim [taVNS] × RewMag [High] × Diff [Hard] | 0.9 | 0.813 | 56.2 | 0.9 | 0.813 | 56.2 | 0.9 | 0.813 | 56.1 |
| Group [MDD] | 0.9 | 0.755 | 55.6 | | | | | | |
| BDI | | | | 0.0 | 0.999 | 55.9 | | | |
| SHAPS | | | | | | | 1.5 | 0.286 | 53.4 |

**Table S7**: Self-rated exertion does not differ between participants with major depressive disorder (MDD) compared to healthy control participants (HCP) in session 1. Results from a linear mixed-effects model**.** BDI = Beck's Depression Inventory. SHAPS = Snaith Hamilton pleasure scale

| | Exertion [Group] | | | Exertion [BDI] | | | Exertion [SHAPS] | | |
|---|---|---|---|---|---|---|---|---|---|
| *Predictors* | *Estimates* | *p* | *df* | *Estimates* | *p* | *df* | *Estimates* | *p* | *df* |
| (Intercept) | 53.0 | **<0.001** | 43.0 | 54.3 | **<0.001** | 35.1 | 54.2 | **<0.001** | 34.2 |
| Stim [taVNS] | 1.0 | 0.833 | 55.9 | 0.8 | 0.854 | 56.2 | 1.0 | 0.828 | 55.9 |
| RewMag [High] | 14.6 | **<0.001** | 28.1 | 14.6 | **<0.001** | 28.2 | 14.6 | **<0.001** | 28.2 |
| Diff [Hard] | -5.1 | **0.027** | 28.0 | -5.1 | **0.027** | 28.0 | -5.1 | **0.027** | 28.0 |
| Money [Money] | 11.0 | **<0.001** | 58.0 | 11.0 | **<0.001** | 58.0 | 11.0 | **<0.001** | 58.0 |
| Stim [taVNS] × RewMag [High] | 6.1 | 0.207 | 52.6 | 6.1 | 0.207 | 52.6 | 6.1 | 0.207 | 52.6 |
| Stim [taVNS] × Diff [Hard] | -7.5 | 0.056 | 51.5 | -7.5 | 0.056 | 51.5 | -7.5 | 0.056 | 51.5 |
| RewMag [High] × Diff [Hard] | 3.0 | 0.161 | 28.0 | 3.0 | 0.161 | 28.5 | 3.0 | 0.161 | 28.2 |
| Stim [taVNS] × RewMag [High] × Diff [Hard] | 2.9 | 0.319 | 56.8 | 2.9 | 0.319 | 57.5 | 2.9 | 0.319 | 57.1 |
| Group [MDD] | 2.4 | 0.410 | 54.4 | | | | | | |
| BDI | | | | 1.6 | 0.255 | 53.5 | | | |
| SHAPS | | | | | | | 1.9 | 0.202 | 55.9 |

**Table S8**: Self-rated exertion does not differ between participants with major depressive disorder (MDD) compared to healthy control participants (HCP) in session 2. Results from a linear mixed-effects model**.** BDI = Beck's Depression Inventory. SHAPS = Snaith Hamilton pleasure scale

| | Invigoration [Group] | | | Invigoration[BDI] | | | Invigoration [SHAPS] | | |
|---|---|---|---|---|---|---|---|---|---|
| *Predictors* | *Estimates* | *p* | *df* | *Estimates* | *p* | *df* | *Estimates* | *p* | *df* |
| (Intercept) | 34.5 | **<0.001** | 103.2 | 34.0 | **<0.001** | 101.7 | 34.0 | **<0.001** | 100.5 |
| Stim [taVNS] | 0.0 | 0.989 | 64.8 | -0.2 | 0.884 | 64.7 | -0.2 | 0.884 | 64.8 |
| RewMag [High] | 6.2 | **0.008** | 59.5 | 8.1 | **<0.001** | 59.0 | 8.1 | **<0.001** | 58.8 |
| Diff [Hard] | 1.7 | 0.319 | 80.0 | 1.1 | 0.353 | 78.0 | 1.1 | 0.352 | 80.5 |
| Session | 4.0 | **<0.001** | 58.4 | 3.9 | **<0.001** | 58.8 | 3.8 | **<0.001** | 60.7 |
| Money [Money] | 5.2 | **<0.001** | 58.0 | 5.2 | **<0.001** | 58.0 | 5.2 | **<0.001** | 58.0 |
| Stim [taVNS] × RewMag[High] | -2.0 | 0.356 | 95.0 | -0.6 | 0.702 | 92.7 | -0.6 | 0.705 | 93.0 |
| Stim [taVNS] × Diff [Hard] | -2.4 | 0.246 | 168.9 | -0.7 | 0.617 | 142.5 | -0.7 | 0.616 | 146.5 |
| RewMag [High] × Diff [Hard] | -0.5 | 0.836 | 68.2 | -1.1 | 0.515 | 68.9 | -1.1 | 0.522 | 67.7 |
| Stim [taVNS] × RewMag [High] × Diff [Hard] | 1.5 | 0.601 | 259.1 | 0.7 | 0.711 | 233.0 | 0.7 | 0.714 | 200.3 |
| Group [MDD] | -1.3 | 0.673 | 57.3 | | | | | | |
| Stim [taVNS] × MDD | -0.4 | 0.871 | 64.9 | | | | | | |
| RewMag [High] × MDD | 3.7 | 0.252 | 59.0 | | | | | | |
| Diff [Hard] × MDD | -1.1 | 0.625 | 80.0 | | | | | | |
| taVNS × RewMag [High] × MDD | 2.8 | 0.357 | 94.8 | | | | | | |
| taVNS × Diff [Hard] × MDD | 3.3 | 0.254 | 174.6 | | | | | | |
| RewMag [High] × Diff [Hard] × MDD | -1.2 | 0.736 | 68.2 | | | | | | |
| taVNS×RewMag [High] × Diff [Hard]) ×Group [MDD] | -1.5 | 0.713 | 264.6 | | | | | | |
| BDI | | | | -0.2 | 0.908 | 57.3 | | | |
| Stim [taVNS] × BDI | | | | -0.5 | 0.739 | 64.8 | | | |

| | Invigoration [Group] | | | Invigoration[BDI] | | | Invigoration [SHAPS] | | |
|---|---|---|---|---|---|---|---|---|---|
| *Predictors* | *Estimates* | *p* | *df* | *Estimates* | *p* | *df* | *Estimates* | *p* | *df* |
| RewMag [High] × BDI | | | | 2.8 | 0.080 | 59.1 | | | |
| Diff [Hard] × BDI | | | | 0.5 | 0.653 | 79.2 | | | |
| taVNS × RewMag [High] × BDI | | | | 1.2 | 0.427 | 93.5 | | | |
| taVNS × Diff [Hard] × BDI | | | | 0.6 | 0.705 | 164.8 | | | |
| RewMag [High] × Diff [Hard] × BDI | | | | -2.5 | 0.131 | 68.8 | | | |
| taVNS × RewMag [High] × Diff [Hard] ×BDI | | | | -0.7 | 0.734 | 253.1 | | | |
| SHAPS | | | | | | | -0.6 | 0.673 | 57.4 |
| Stim [taVNS] × SHAPS | | | | | | | 0.9 | 0.519 | 64.8 |
| RewMag [High] × SHAPS | | | | | | | 1.2 | 0.443 | 58.9 |
| Diff [Hard] × SHAPS | | | | | | | 1.1 | 0.353 | 82.0 |
| taVNS × RewMag [High] × SHAPS | | | | | | | 2.0 | 0.209 | 94.0 |
| taVNS × Diff [Hard]) × SHAPS | | | | | | | 1.3 | 0.388 | 169.5 |
| RewMag [High] × Diff [Hard] × SHAPS | | | | | | | -1.1 | 0.516 | 67.7 |
| taVNS × RewMag [High] × Diff [Hard]) ×SHAPS | | | | | | | -1.9 | 0.342 | 212.8 |

**Table S9**: Invigoration slope is not altered by transcutaneous vagus nerve stimulation (taVNS) across both sessions and independent of group. Results from a linear mixed effects models**.** BDI = Beck’s Depression Inventory. SHAPS = Snaith Hamilton pleasure scale

| | Effort maintenance [Group] | | | Effort maintenance [SHAPS] | | | Effort maintenance [BDI] | | |
|---|---|---|---|---|---|---|---|---|---|
| *Predictors* | *Estimates* | *p* | *df* | *Estimates* | *p* | *df* | *Estimates* | *p* | *df* |
| (Intercept) | 49.4 | **<0.001** | 79.4 | 58.9 | **<0.001** | 59.7 | 60.0 | **<0.001** | 59.8 |
| Stim [taVNS] | 1.3 | 0.598 | 56.8 | 0.8 | 0.647 | 56.8 | 0.8 | 0.648 | 56.8 |
| RewMag High] | 9.5 | **0.002** | 58.1 | 10.6 | **<0.001** | 57.2 | 10.6 | **<0.001** | 57.2 |
| Diff [Hard] | -9.2 | **<0.001** | 57.4 | -8.7 | **<0.001** | 57.0 | -8.7 | **<0.001** | 57.1 |
| Session | 3.6 | **<0.001** | 62.3 | 3.7 | **<0.001** | 60.0 | 3.6 | **<0.001** | 60.4 |
| Money [Money] | 7.8 | **<0.001** | 58.0 | 7.8 | **<0.001** | 58.1 | 7.8 | **<0.001** | 58.0 |
| Stim [taVNS] × RewMag [High] | -1.2 | 0.636 | 58.3 | -0.5 | 0.757 | 58.3 | -0.5 | 0.757 | 58.3 |
| Stim [taVNS] × Diff [Hard] | -2.1 | 0.237 | 57.3 | -2.1 | 0.104 | 57.3 | -2.1 | 0.104 | 57.3 |
| RewMag [High] × Diff [Hard] | 3.5 | **0.026** | 65.9 | 3.4 | **0.003** | 66.0 | 3.4 | **0.003** | 66.0 |
| Stim [taVNS] × RewMag [High] × Diff [Hard] | -0.3 | 0.889 | 74.5 | 1.5 | 0.332 | 73.1 | 1.5 | 0.331 | 73.4 |
| Group [MDD] | 3.0 | 0.519 | 57.0 | | | | | | |
| Stim [taVNS] × Group [MDD] | -1.0 | 0.770 | 56.8 | | | | | | |
| RewMag [High] × Group [MDD] | 2.3 | 0.572 | 57.1 | | | | | | |
| Diff [Hard] × Group [MDD] | 1.0 | 0.749 | 57.0 | | | | | | |
| Stim [taVNS] × RewMag [High] × Group [MDD] | 1.2 | 0.719 | 58.3 | | | | | | |
| Stim [taVNS] × Diff [Hard] × Group [MDD] | 0.1 | 0.959 | 57.3 | | | | | | |
| RewMag [High] × Diff [Hard] × Group [MDD] | -0.2 | 0.917 | 65.8 | | | | | | |
| Stim [taVNS] × RewMag [High] × Diff [Hard] × Group [MDD] | 3.5 | 0.248 | 74.3 | | | | | | |
| SHAPS | | | | -0.9 | 0.675 | 53.7 | | | |

| | | | | | | |
|---|---|---|---|---|---|---|
| Stim [taVNS] × SHAPS | 1.4 | 0.408 | 56.8 | | | |
| RewMag [High] × SHAPS | 0.5 | 0.805 | 57.2 | | | |
| Diff [Hard] × SHAPS | -0.5 | 0.763 | 57.0 | | | |
| Stim [taVNS] × RewMag [High] × SHAPS | -0.3 | 0.884 | 58.3 | | | |
| Stim [taVNS] × Diff [Hard] × SHAPS | 0.4 | 0.774 | 57.3 | | | |
| RewMag [High] × Diff [Hard] × SHAPS | -0.3 | 0.784 | 66.0 | | | |
| Stim [taVNS] × RewMag [High] × Diff [Hard] × SHAPS | 0.9 | 0.528 | 73.5 | | | |
| BDI | | | | 1.1 | 0.616 | 55.8 |
| Stim [taVNS] × BDI | | | | -0.9 | 0.602 | 56.9 |
| RewMag [High] × BDI | | | | 0.5 | 0.790 | 57.1 |
| Diff [Hard] × BDI | | | | 1.4 | 0.396 | 57.0 |
| Stim [taVNS] × RewMag [High] × BDI | | | | 1.0 | 0.561 | 58.3 |
| Stim [taVNS] × Diff [Hard] × BDI | | | | -0.0 | 0.969 | 57.3 |
| RewMag [High] × Diff [Hard] × BDI | | | | -0.1 | 0.924 | 66.0 |
| Stim [taVNS] × RewMag [High] × Diff [Hard] × BDI | | | | 1.3 | 0.397 | 73.8 |

**Table S10**: Effort maintenance is not altered by transcutaneous vagus nerve stimulation (taVNS) across both sessions and independent of group. Results from a linear mixed effects models**.** BDI = Beck's Depression Inventory. SHAPS = Snaith Hamilton pleasure scale

| | Wanting [Group] | | | Wanting [BDI] | | | Wanting [SHAPS] | | |
|---|---|---|---|---|---|---|---|---|---|
| *Predictors* | *Estimates* | *p* | *df* | *Estimates* | *p* | *df* | *Estimates* | *p* | *df* |
| (Intercept) | 49.4 | **<0.001** | 89.1 | 45.2 | **<0.001** | 87.2 | 45.3 | **<0.001** | 87.8 |
| Stim [taVNS] | -0.3 | 0.910 | 55.6 | -1.3 | 0.466 | 54.3 | -1.3 | 0.465 | 54.5 |
| RewMag [High] | 17.6 | **<0.001** | 60.3 | 21.0 | **<0.001** | 57.2 | 21.0 | **<0.001** | 57.4 |
| Diff [Hard] | -7.8 | **0.001** | 57.7 | -7.6 | **<0.001** | 57.9 | -7.6 | **<0.001** | 57.8 |
| Session | 3.1 | **0.003** | 56.2 | 3.2 | **0.003** | 56.2 | 3.2 | **0.003** | 56.2 |
| Money [Money] | 15.3 | **<0.001** | 58.0 | 15.3 | **<0.001** | 58.0 | 15.3 | **<0.001** | 58.0 |
| Stim [taVNS] × RewMag [High] | -0.8 | 0.777 | 60.2 | 0.1 | 0.960 | 59.1 | 0.1 | 0.960 | 59.1 |
| Stim [taVNS] × Diff [Hard] | 1.0 | 0.567 | 86.4 | 0.3 | 0.831 | 115.9 | 0.3 | 0.833 | 85.3 |
| RewMag [High] × Diff [Hard] | 4.2 | **0.031** | 79.8 | 3.7 | **0.007** | 81.3 | 3.7 | **0.008** | 80.6 |
| Stim [taVNS] × RewMag [High] × Diff [Hard] | 0.4 | 0.865 | 179.8 | 0.8 | 0.612 | 208.1 | 0.8 | 0.617 | 166.2 |
| Group [MDD] | -8.0 | 0.087 | 57.0 | | | | | | |
| Diff [Hard] × Group [MDD] | 0.4 | 0.908 | 57.7 | | | | | | |
| taVNS × MDD | -2.0 | 0.567 | 54.8 | | | | | | |
| RewMag [High] × MDD | 6.8 | 0.131 | 57.3 | | | | | | |
| taVNS × RewMag [High] × MDD] | 1.8 | 0.652 | 59.2 | | | | | | |
| taVNS × Diff [Hard] × MDD | -1.5 | 0.549 | 86.1 | | | | | | |
| RewMag [High] × Diff [Hard] × MDD | -1.1 | 0.684 | 79.7 | | | | | | |
| taVNS× RewMag [High] × Diff [Hard]) × MDD | 0.8 | 0.797 | 183.0 | | | | | | |
| BDI | | | | -2.8 | 0.239 | 57.0 | | | |

| | Wanting [Group] | | | Wanting [BDI] | | | Wanting [SHAPS] | | |
|---|---|---|---|---|---|---|---|---|---|
| *Predictors* | *Estimates* | *p* | *df* | *Estimates* | *p* | *df* | *Estimates* | *p* | *df* |
| taVNS × BDI | | | | -0.1 | 0.975 | 54.8 | | | |
| RewMag [High] × BDI | | | | 4.1 | 0.068 | 57.3 | | | |
| Diff [Hard] × BDI | | | | 1.8 | 0.246 | 57.9 | | | |
| taVNS × RewMag [High] × BDI | | | | -0.8 | 0.675 | 59.3 | | | |
| taVNS × Diff [Hard]) × BDI | | | | -2.1 | 0.080 | 118.8 | | | |
| RewMag [High] × Diff [Hard]) × BDI | | | | -1.8 | 0.171 | 81.7 | | | |
| taVNS × RewMag[High] × Diff [Hard] × BDI | | | | 2.5 | 0.114 | 232.8 | | | |
| SHAPS | | | | | | | -3.4 | 0.150 | 57.0 |
| taVNS × SHAPS | | | | | | | 0.0 | 0.985 | 54.7 |
| RewMag [High] × SHAPS | | | | | | | 3.5 | 0.121 | 57.3 |
| Diff [Hard] × SHAPS | | | | | | | 2.0 | 0.183 | 57.8 |
| taVNS × RewMag [High] × SHAPS | | | | | | | -1.2 | 0.559 | 59.2 |
| taVNS × Diff [Hard] × SHAPS | | | | | | | -1.4 | 0.250 | 86.7 |
| RewMag [High] × Diff [Hard] × SHAPS | | | | | | | -1.6 | 0.240 | 81.1 |
| taVNS × RewMag [High] × Diff [Hard] × SHAPS | | | | | | | 0.7 | 0.663 | 185.9 |

**Table S11**: Wanting is not altered by transcutaneous vagus nerve stimulation (taVNS) across both sessions and independent of group. Results from a linear mixed effects models**.** BDI = Beck's Depression Inventory. SHAPS = Snaith Hamilton pleasure scale
